# Large language models outperform traditional structured data-based approaches in identifying immunosuppressed patients

**DOI:** 10.1101/2025.01.16.25320564

**Authors:** Vijeeth Guggilla, Mengjia Kang, Melissa J Bak, Steven D Tran, Anna Pawlowski, Prasanth Nannapaneni, Luke V Rasmussen, Daniel Schneider, Helen Donnelly, Ankit Agrawal, David Liebovitz, Alexander V Misharin, GR Scott Budinger, Richard G Wunderink, Theresa L Walunas, Catherine A Gao, The NU SCRIPT Study Investigators

## Abstract

Identifying immunosuppressed patients using structured data can be challenging. Large language models effectively extract structured concepts from unstructured clinical text. Here we show that GPT-4o outperforms traditional approaches in identifying immunosuppressive conditions and medication use by processing hospital admission notes. We also demonstrate the extensibility of our approach in an external dataset. Cost-effective models like GPT-4o mini and Llama 3.1 also perform well, but not as well as GPT-4o.

## Introduction

Immunosuppressed individuals are at heightened risk of morbidity and mortality in the context of infectious disease and especially pneumonia, which is responsible for over 40,000 deaths every year in the United States alone^1^. Not only do many pre-existing conditions cause immunosuppression but numerous immunosuppressive medications can also significantly increase patient vulnerability to infection. While structured data-based approaches of identifying immunosuppression using diagnosis codes and medication orders from the electronic health record (EHR) have been developed, such strategies tend to have limited accuracy and are difficult to extend to multiple heterogeneous types of immunosuppression^2^.

Clinical notes are a rich source of patient information and can be useful in building patient cohorts to answer specific clinical or research questions. However, because clinical notes are unstructured, significant effort is required to parse information from them in high throughput contexts^3^. While many natural language processing (NLP) approaches have been developed to automate data extraction from clinical notes, large language models (LLM) stand out in their ability to work with unstructured text, exhibiting human-level performance on NLP benchmarks across domains^4–6^. LLMs have impressively replicated clinical expertise across multiple medical specialties^7–9^. For example, LLMs can effectively extract structured concepts from pathology reports and identify obsessive-compulsive disorder from medical vignettes^10,11^. Given LLMs’ rapidly expanding potential to reduce human effort associated with processing clinical text, testing their application in different medical contexts for added value is important.

Using LLMs to extract information about immunosuppression from clinical notes is a potential alternative to relying on structured data and manual curation. As LLMs have not yet been tested in this important task, our objective was to evaluate how LLMs would perform in identifying patients with immunosuppressive conditions or taking immunosuppressive medications by analyzing the text of hospital admission notes. We compared LLM results to traditional diagnosis code-based and medication order-based approaches of identifying immunosuppression.

## Results

We identified 827 admissions at Northwestern Memorial Hospital between June 2018 and August 2024 for patients who were mechanically ventilated for suspected pneumonia to test the performance of structured data and LLMs in identifying immunosuppressive conditions and medication use (Fig. 1). For diagnosis code-based identification, we extracted all diagnosis codes a patient received prior to hospitalization and coded conditions as positive based on the number of dates a patient received a certain code. F1 scores varied across different frequency thresholds for each condition. For example, the F1 score for identifying multiple myeloma was 0.71 at a threshold of n = 1 date on which a multiple myeloma diagnosis code was present. However, the best F1 score for multiple myeloma was 0.97 at a threshold of n = 5 dates. Peak F1 scores were 0.83 or above for all clinical conditions except immunoglobulin deficiency, for which the F1 score was 0.48 (Fig. 2A) (Supplementary tables 1, 2). We tested how the GPT-4o LLM would perform given hospital admission notes and an LLM prompt modeled after a previously published prompt that successfully extracted structured concepts from pathology notes^10^ (Fig. 3). F1 scores were extremely high overall and outperformed structured data for all conditions: 0.99 for acute leukemia, 1 for HIV, 0.51 for immunoglobulin deficiency, 0.96 for lymphoma, 0.97 for multiple myeloma, 0.97 for solid organ transplant, and 0.99 for stem cell transplant (Fig. 4A) (Supplementary tables 1, 3). Because the prompt asks the LLM to not only classify each variable but also to provide its reasoning, we were able to better understand the false positives and negatives. For example, many false positive immunoglobulin deficiency classifications were due to the LLM mistakenly reasoning that intravenous immunoglobulin administration must coincide with an immunoglobulin deficiency. False negatives were rare and were often due to admission notes missing information about patients’ complete medical histories.

**Fig. 1:**
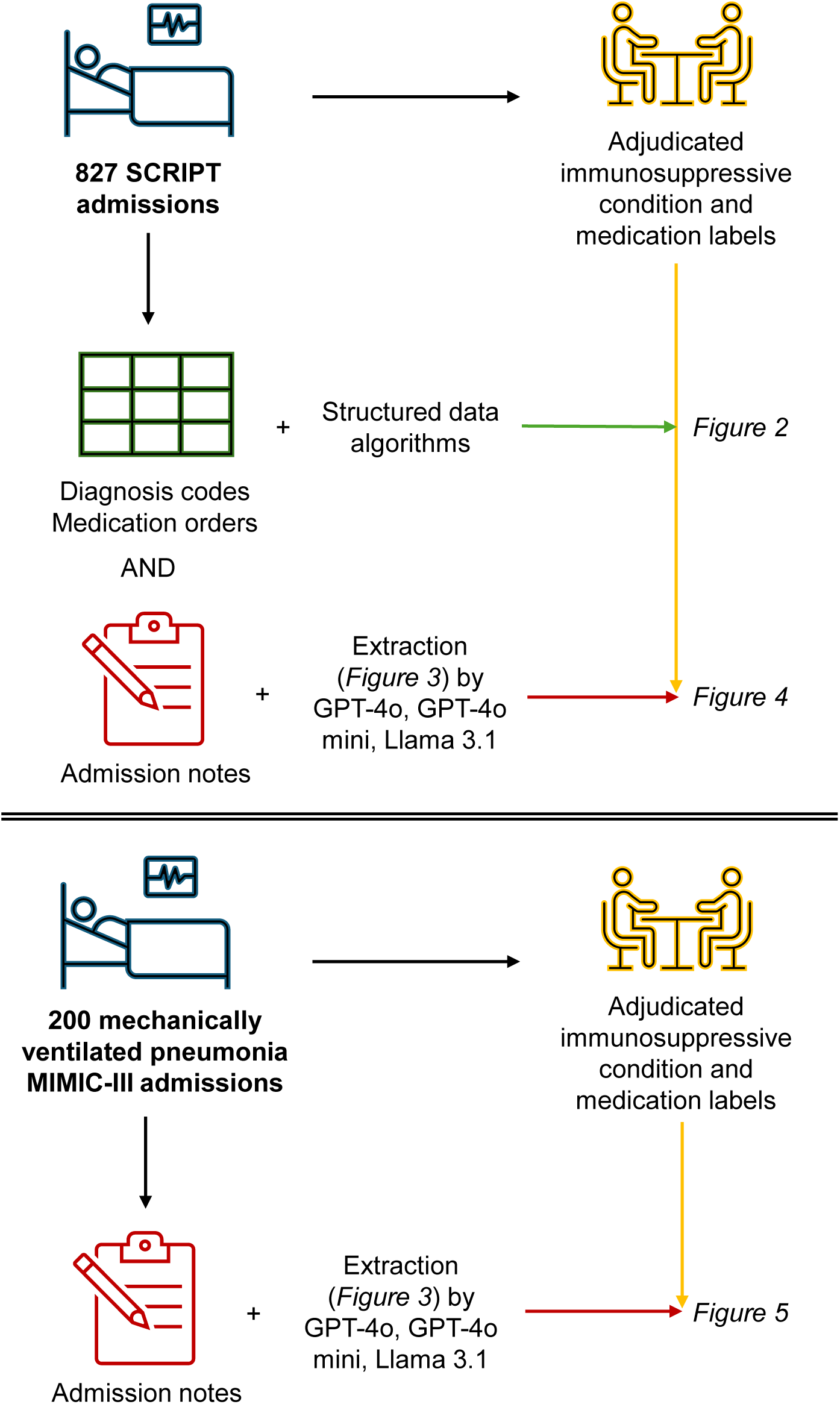
Flow diagram of study design comparing structured data and LLM performance in identifying immunosuppression followed by external validation of LLM performance. Structured data and admission notes were extracted for 827 SCRIPT admissions which were adjudicated for immunosuppressive conditions and medications. The performance of structured data algorithms and LLMs in predicting the adjudicated labels was assessed. External validation of using LLMs to predict adjudicated immunosuppression labels was performed in 200 MIMIC-III admission notes.

**Fig. 2:**
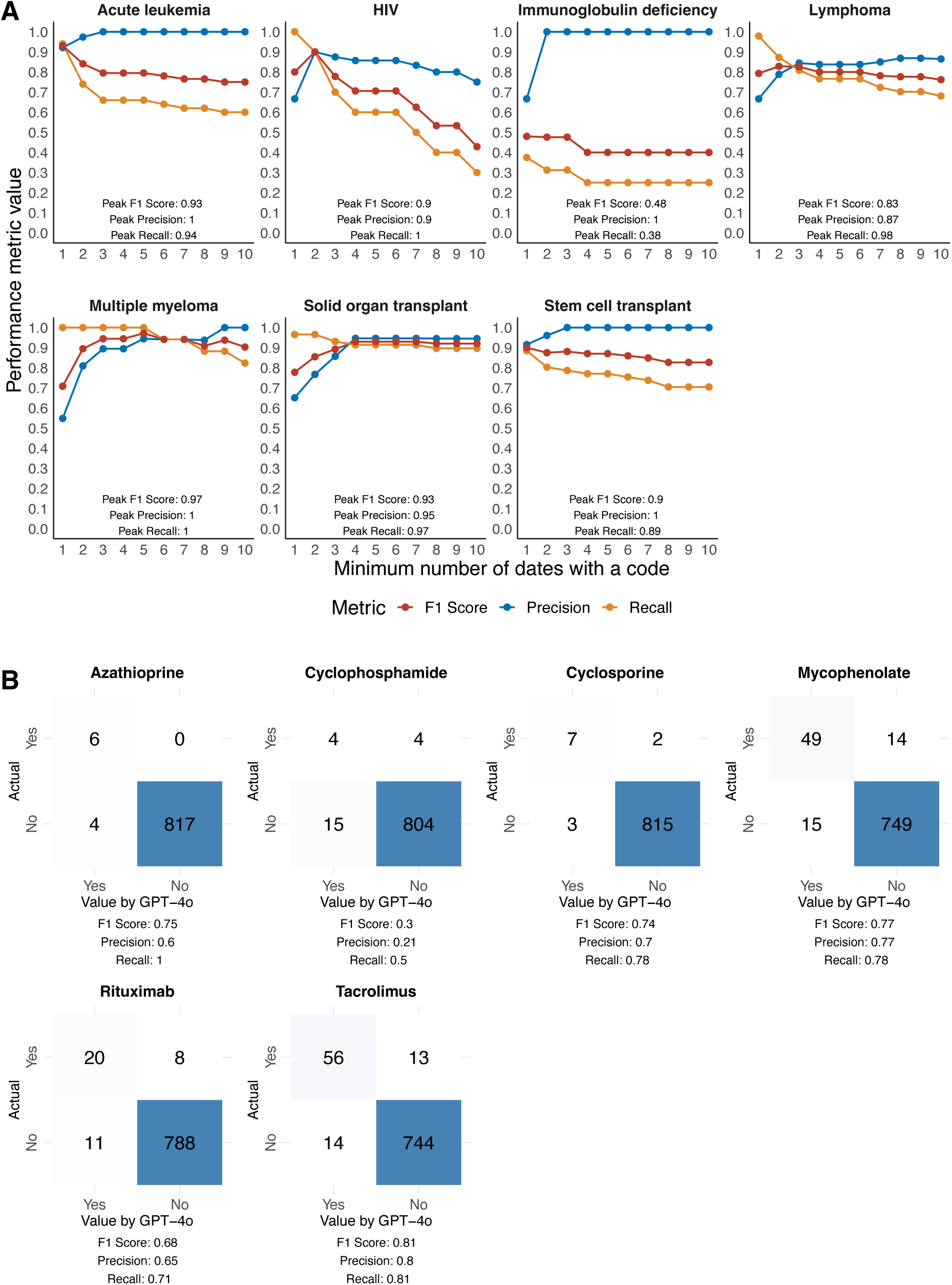
Performance of structured data in identifying immunosuppressive conditions and medications. F1 scores, precision, and recall were computed by comparing structured data predictions to adjudicated labels. **A** Performance metrics across an increasing minimum number of dates with a diagnosis code required to count as a case. **B** Confusion matrices for predicting medication use by the presence of a valid medication order in the six months prior to admission.

**Fig. 3:**
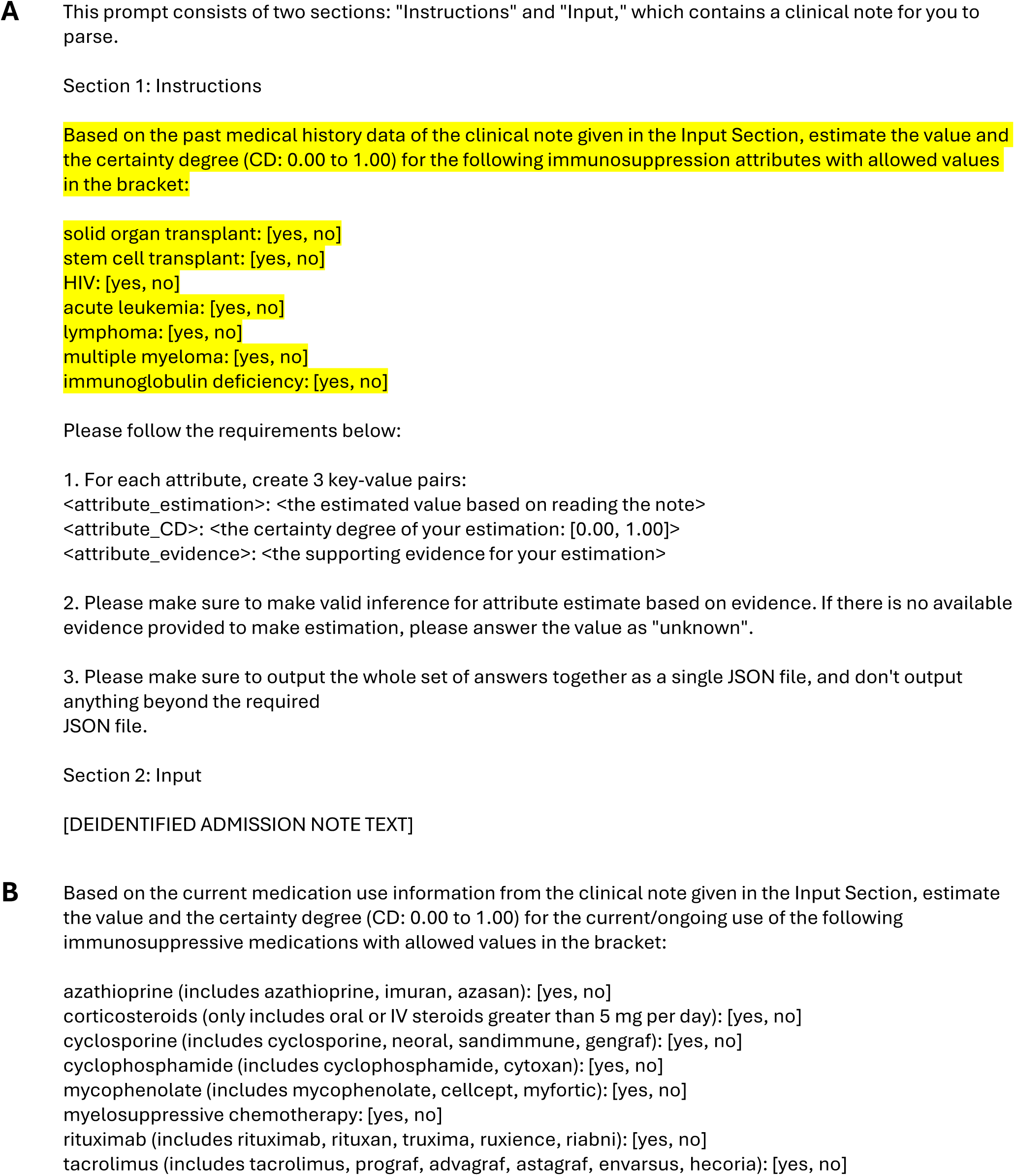
Prompts used for immunosuppression attribute extraction and estimation from admission notes. Prompts for extracting **A** immunosuppressive conditions and **B** immunosuppressive medications where the highlighted portion is either a list of **A** conditions or **B** medications.

**Fig. 4:**
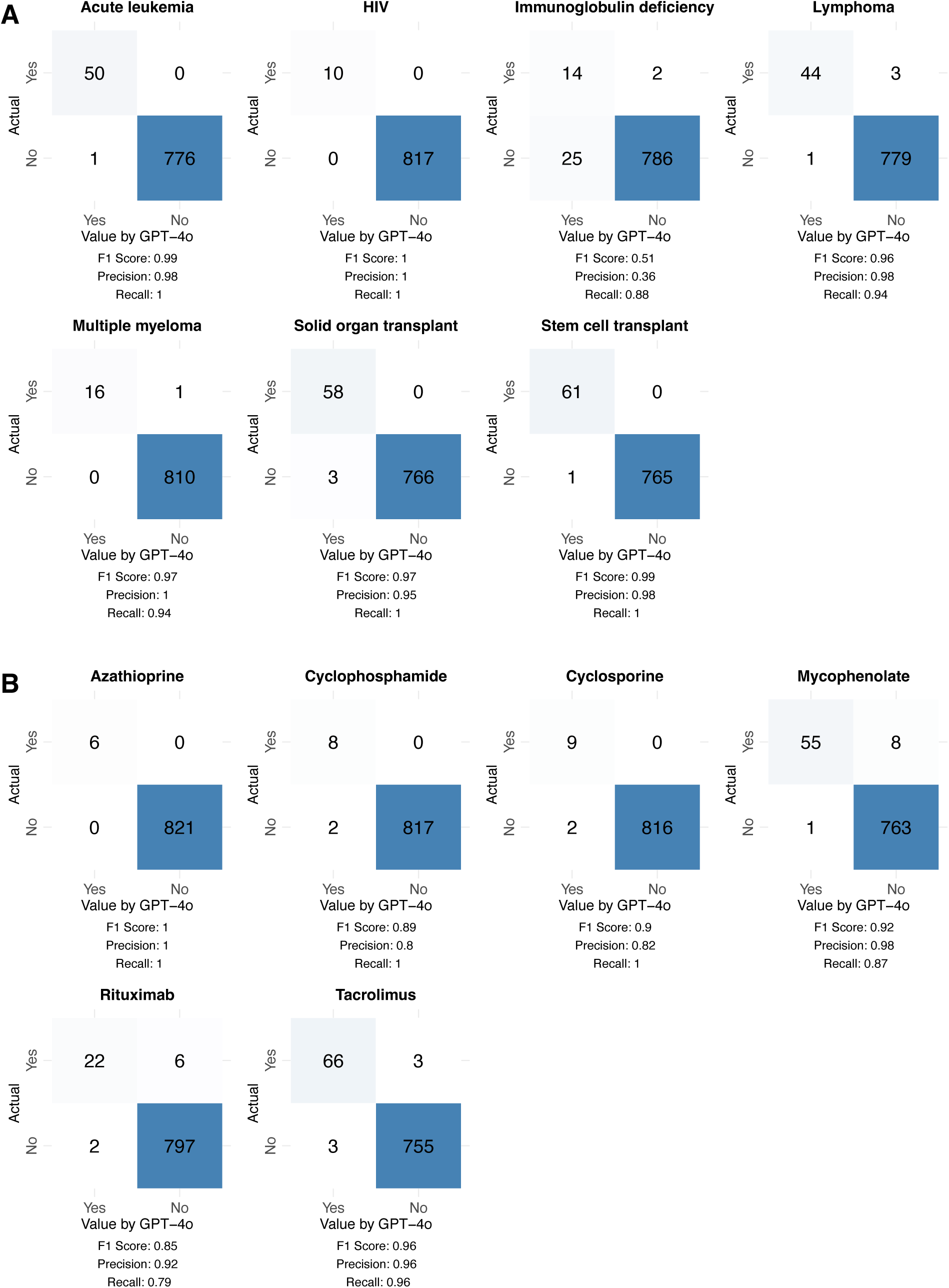
Performance of GPT-4o in identifying immunosuppressive conditions and medications in the SCRIPT cohort. F1 scores, precision, and recall were computed by comparing GPT-4o predictions to adjudicated labels. Confusion matrices for GPT-4o predicting **A** immunosuppressive conditions and **B** immunosuppressive medication use from admission notes.

We also tested how medication orders would perform in identifying patients who are on various immunosuppressive medications. F1 scores were 0.68 or above for all medications except cyclophosphamide, for which the F1 score was 0.3 (Fig. 2B) (Supplementary tables 1, 2). The GPT-4o LLM performed more consistently and outperformed structured data for every medication, with F1 scores of 1 for azathioprine, 0.89 for cyclophosphamide, 0.9 for cyclosporine, 0.92 for mycophenolate, 0.85 for rituximab, and 0.96 for tacrolimus (Fig. 4B) (Supplementary tables 1, 3). Most false positives were due to newly ordered medications being misinterpreted as medications that were already being taken prior to admission, which could be related to the wording “current/ongoing use” in our prompt. False negatives were either due to recent holds on medications being misinterpreted as a patient never having been on a medication or due to notes missing information about medication use.

To support use of LLMs in environments with lower computational resources or only permission to use locally-run LLMs, we also wanted to test how simpler models would perform at these tasks. We first tested GPT-4o mini, a cost-efficient GPT-4o alternative^12^. GPT-4o mini achieved F1 scores ranging from 0.68 to 0.97 for immunosuppressive conditions, and F1 scores ranging from 0.56 to 0.94 for immunosuppressive medications (Supplementary tables 1, 4). We also tested Llama 3.1, a cost-efficient open-source model that can run locally^13^. Llama 3.1 achieved F1 scores ranging from 0.26 to 0.93 for immunosuppressive conditions, and F1 scores ranging from 0.09 to 0.89 for immunosuppressive medications (Supplementary table 1, 5).

External validation is important when developing EHR-based phenotyping approaches to assess the generalizability of proposed methods across healthcare institutions and the robustness of said methods against site-specific artefacts. We performed external validation in the Medical Information Mart for Intensive Care III (MIMIC-III) dataset, which contains health-related data and clinical notes associated with patients who stayed in critical care units at Beth Israel Deaconess Medical Center between 2001 and 2012^14–16^. We identified 200 admissions for patients in MIMIC-III who were mechanically ventilated and admitted with a diagnosis of pneumonia who also had admission notes available. We tested GPT-4o, GPT-4o mini, and Llama 3.1 with the same prompts in the MIMIC-III corpus we developed. Our corpus had true positives present for every condition and medication of interest except immunoglobulin deficiency and azathioprine. GPT-4o performed extremely well across the board, with F1 scores of 1 for many conditions and medications. The only variables with poor performance were immunoglobulin deficiency and cyclophosphamide, for which the F1 scores were 0 (Fig. 5) (Supplementary tables 1, 6). GPT-4o mini achieved F1 scores ranging from 0.86 to 1 for immunosuppressive conditions other than immunoglobulin deficiency. For medications other than cyclophosphamide, F1 scores ranged from 0.67 to 1 (Supplementary tables 1, 7). Llama 3.1 achieved F1 scores ranging from 0.15 to 0.89 for immunosuppressive conditions other than immunoglobulin deficiency. For medications, F1 scores ranged from 0 to 0.8 (Supplementary tables 1, 8).

**Fig. 5:**
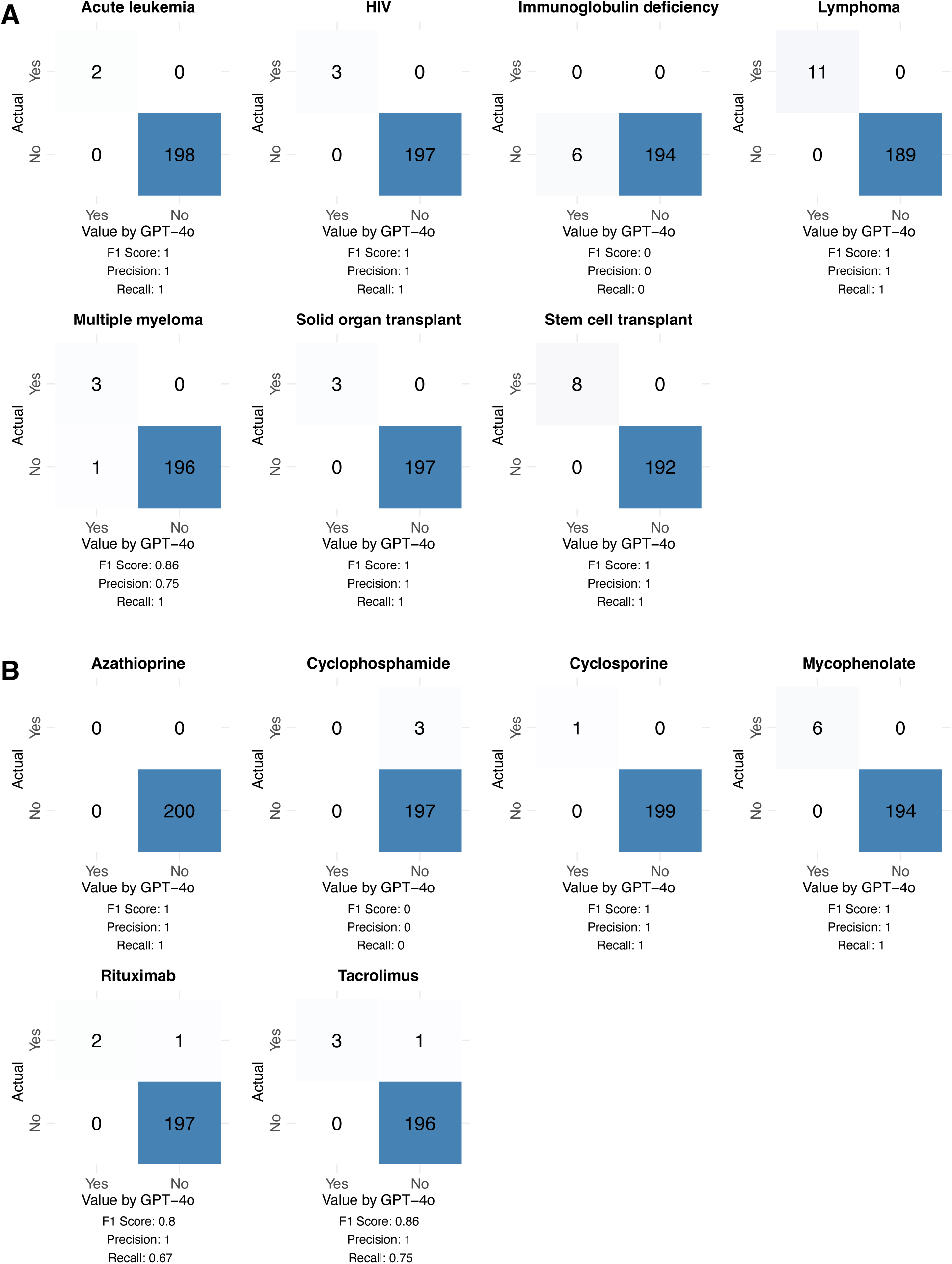
Performance of GPT-4o in identifying immunosuppressive conditions and medications in the MIMIC-III cohort. F1 scores, precision, and recall were computed by comparing GPT-4o predictions to adjudicated labels. Confusion matrices for GPT-4o predicting **A** immunosuppressive conditions and **B** immunosuppressive medication use from admission notes.

## Discussion

LLMs can effectively extract history of immunosuppression from clinical notes. For every immunosuppressive condition and medication examined, GPT-4o outperformed a rules-based structured data approach, achieving F1 scores above 0.84 for all variables except immunoglobulin deficiency, for which performance suffered mainly due to its vague definition in the prompt. GPT-4o mini also performed well but suffered from more false positives than GPT-4o. Many of these false positives were due to GPT-4o mini misinterpreting risks of or evaluations for certain conditions as the presence of those conditions. For example, patients who were evaluated for a solid organ transplant were misclassified as patients who had received a solid organ transplant. Llama 3.1 was the weakest performer, but it did exhibit impressive performance for certain variables such as stem cell transplant, lymphoma, tacrolimus, and mycophenolate. Llama 3.1 was even more prone to the type of false positives GPT-4o mini suffered from, mistaking possibilities of certain conditions as evidence of these conditions. For example, Llama 3.1 mistakenly reasoned that patients who may require surgical intervention had received a solid organ transplant. It also made simpler mistakes such as confusing azathioprine and azithromycin.

One limitation of the cohorts we identified is that they consisted of critically ill patients. Our approach has not been tested in cohorts of patients who were not necessarily hospitalized and may have differently detailed notes available for LLMs to process. In general, this approach is intrinsically limited by available note quality. Another limitation of using LLMs like GPT-4o is their cost. For example, processing either conditions or medications with GPT-4o using a full admission note costs approximately 5 cents per note. Thus, the cost of processing conditions for all 827 notes in our SCRIPT cohort was about $50. While 5 cents per note is relatively inexpensive, it is important to keep in mind when considering scaling our approach. GPT-4o mini, on the other hand, only cost approximately 0.1 cent per note.

Overall, our results show promise for leveraging LLMs to accurately extract structured information about immunosuppression from unstructured clinical notes. Just as we have expanded this framework from pathology notes to clinical notes, we hope that these approaches can be extended toward other types of notes or information to further improve cohort identification and adjudication. To this end, we share our prompts, code repositories, and datasets for the scientific community’s endeavors.

## Methods

### Data source

We utilized data from patients enrolled in Successful Clinical Response in Pneumonia Therapy (SCRIPT), a single-center prospective cohort study (IRB STU00204868) of patients at Northwestern Memorial Hospital who required mechanical ventilation and underwent a bronchoalveolar lavage for suspected pneumonia between June 2018 and August 2024^17^. The patient cohort was 59% male, 59% White, 19% Black, 19% Hispanic, and had a mean age of 60 years. All cause hospital mortality was 41%. The research team manually reviewed each patient chart at time of study enrollment for the presence of the following pre-existing immunosuppressive conditions: acute leukemia, HIV, immunoglobulin deficiency, lymphoma, multiple myeloma, solid organ transplant, and stem cell transplant. They also reviewed for a history of using the following immunosuppressive medications: azathioprine, cyclosporine, cyclophosphamide, mycophenolate, rituximab, and tacrolimus^18^. V.G. re-reviewed 139 cases where there was uncertainty.

### Structured data performance

For each patient, all International Classification of Diseases 9 and 10 (ICD-9 and ICD-10) diagnosis codes accumulated in their medical record prior to hospitalization and admission notes corresponding to the given hospitalization were extracted from the Northwestern Medicine Enterprise Data Warehouse. Patients were categorized as having the immunosuppressive conditions listed above based on the presence of ICD-9 or ICD-10 codes corresponding to that condition on *n* or more different dates before admission (Supplementary table 9). Precision, recall, and F1 score for identification of each immunosuppressive condition compared to the gold standard definition from chart review were computed at values of *n* from one to ten. In addition, for each patient, medication order data for six months prior to hospitalization was extracted from the EHR. The presence of a valid order for a given medication was used to categorize patients as a user of that medication (Supplementary table 9). Precision, recall, and F1 score for identification of each immunosuppressive medication compared to the gold standard definition from chart review were computed.

### LLM performance

We aimed to compare how automated extraction of these immunosuppressive conditions and medications from clinical admission notes using an LLM would perform. Admission notes were first processed through *Philter*, an open-source clinical text de-identification tool, to remove protected health information^19^. Microsoft Azure’s secure OpenAI API for batch querying gpt-4o-2024-02-01 (GPT-4o) was used to analyze the notes and extract immunosuppressive conditions. For each note, GPT-4o was given a prompt modeled after a previously published prompt that successfully extracted concepts from clinical text along with the entire deidentified note text^10^. Hyperparameters were set as follows: temperature = 0, max tokens = 4000, probability mass = 1, presence penalty = 0, and frequency penalty = 0. The prompt instructed GPT-4o to output immunosuppression labels and evidence for each classification in JSON format for simplified post-processing. The prompt also instructed the LLM to output its certainty of each classification to encourage ‘chain of thought’ reasoning^20^. Precision, recall, and F1 score for identification of each immunosuppressive condition and medication compared to the gold standard definition from chart review were computed. LLM analyses were also repeated with GPT-4o mini and Llama 3.1 using the same prompt and parameters that were used with GPT-4o. GPT-4o mini was accessed through Microsoft Azure’s secure OpenAI API for batch querying gpt-4o-mini-2024-02-01. Llama 3.1 was downloaded from bartowski/Meta-Llama-3.1-8B-Instruct-GGUF/Meta-Llama-3.1-8B-Instruct-Q8_0.gguf on LM Studio version 0.2.31 and run locally^21^.

### External validation

We performed external validation of our LLM-based approaches in MIMIC-III, a clinical database of patients who stayed in critical care units at Beth Israel Deaconess Medical Center between 2001 and 2012^14–16^. The MIMIC-III ventilation table was used to filter for patients who had been mechanically ventilated and admitted to the medical intensive care unit. Diagnosis codes were used to further filter for patients who had been admitted with pneumonia. Finally, patients were filtered for those with admission notes available. Once our MIMIC-III cohort and associated corpus of admission notes was identified, each note was manually adjudicated for the presence or absence of the same immunosuppressive conditions and medications described above. Each note was reviewed by at least two independent reviewers, with a third tiebreaker reviewer for disagreements or unsure categorizations. Using this corpus, we repeated the same analyses described above with GPT-4o, GPT-4o-mini, and Llama 3.1.

All analyses were performed in Python 3.12.4. Complete analysis code is available at https://github.com/NUSCRIPT/guggilla_immunosuppression_2025.

## Data availability

The annotated SCRIPT notes dataset analyzed during the current study is available from the corresponding author upon reasonable request to those who sign a Data Use Agreement. MIMIC-III is available at https://physionet.org/content/mimiciii/1.4/ to credentialed users. The annotated MIMIC-III notes corpus developed and analyzed during the current study has been submitted to PhysioNet as a derived dataset to make it readily available to credentialed users of MIMIC-III. In the meanwhile, it is available to credentialed users from the corresponding author upon reasonable request.

## Code availability

The underlying code for this study is available in the repository “guggilla_immunosuppression_2025” and can be accessed via this link: https://github.com/NUSCRIPT/guggilla_immunosuppression_2025. The code is written in Python 3.12.4.

## Acknowledgements

The NU SCRIPT Study is funded by NIH NIAID U19AI135964. This work was also supported by NUCATS, SQLIFTS, and the Canning Thoracic Institute of Northwestern Medicine. S.D.T. was supported by the NIH (grant no. 1F31LM014201). A.A. was supported by NIH (grant nos. U19AI135964 and R01HL158139). A.V.M. was supported by the NIH (grant nos. U19AI135964, P01AG049665, P01HL154998, U19AI181102, R01HL153312, R01HL158139, R01ES034350, and R21AG075423). G.R.S.B. was supported by a Chicago Biomedical Consortium grant, a Northwestern University Dixon Translational Science Award, the Simpson Querrey Lung Institute for Translational Science, the NIH (grant nos. P01AG049665, P01HL154998, U54AG079754, R01HL147575, R01HL158139, R01HL147290, R21AG075423, and U19AI135964), and the Veterans Administration (award no. I01CX001777). R.G.W. was supported by the NIH (grant nos. U19AI135964, U01TR003528, P01HL154998, R01HL149883, R01LM013337). T.L.W was supported by Gilead Sciences (award no. CO-US-540-6435) and the NIH (grant nos. U19AI135964, U19AI181102, and R21HD107571). C.A.G was supported by the NIH (grant no. K23HL169815), a Parker B. Francis Opportunity Award, and an ATS Unrestricted Grant.

## Author contributions

V.G. was involved in study design, methodology development, data acquisition, experimental analysis and visualization, drafting the manuscript, and chart review. M.K. was involved in methodology development, experimental analysis and visualization, and chart review. M.B. was involved in chart review. S.D.T. was involved in experimental analysis and visualization. A.P., P.N., L.R., D.S., and H.D. were involved in data acquisition. A.A. and D.L. were involved in methodology development and study supervision. A.V.M, G.R.S.B., and R.G.W. were involved in funding acquisition and study supervision. T.L.W. was involved in study design, methodology development, funding acquisition, and study supervision. C.A.G. was involved in study conception, study design, methodology development, experimental analysis and visualization, chart review, and study supervision. All authors were involved in reviewing and editing, and all authors have read and approved the final manuscript.

## Ethics declarations

### Competing interests

T.L.W. has received research funding from Gilead Sciences to support investigation of the relationship between immunosuppressive conditions and COVID-19 outcomes. Gilead personnel had no involvement in this research. All other authors declare no financial or non-financial competing interests.

## Supplementary material

**Supplementary table 1.**
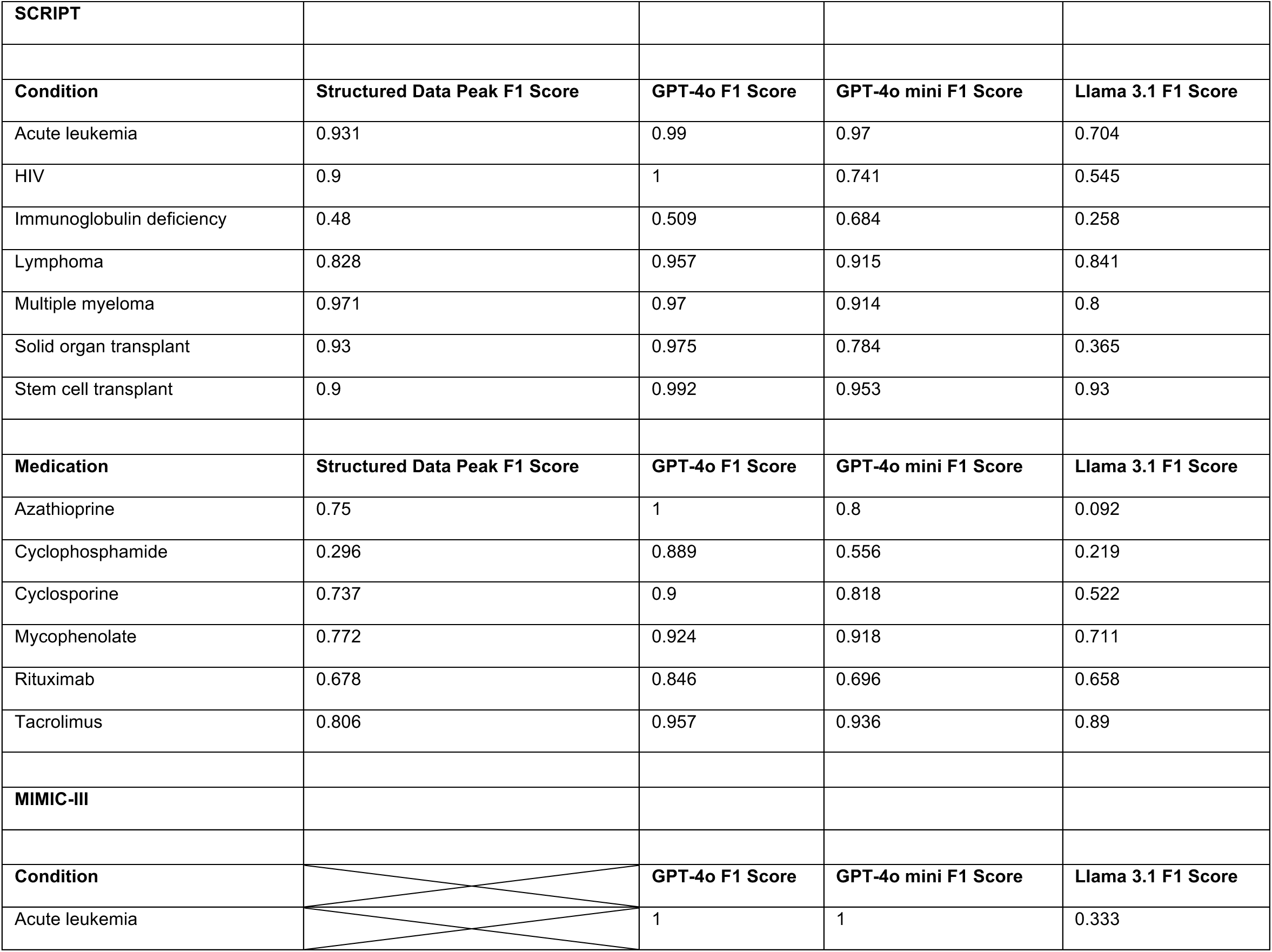
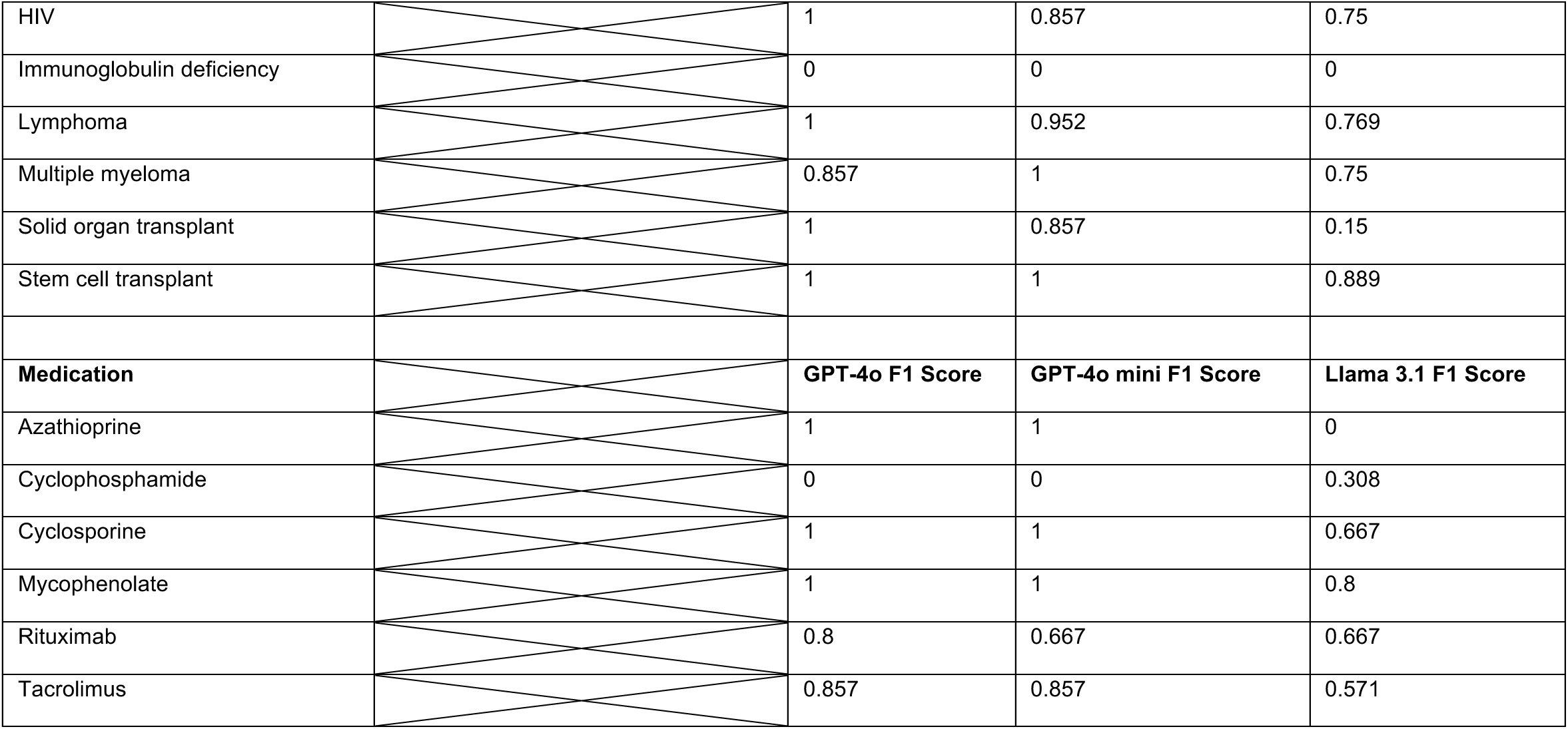
Summary of F1 scores for all identification methods of all immunosuppressive conditions and medications in both the SCRIPT and MIMIC-III cohorts.

**Supplementary table 2.**
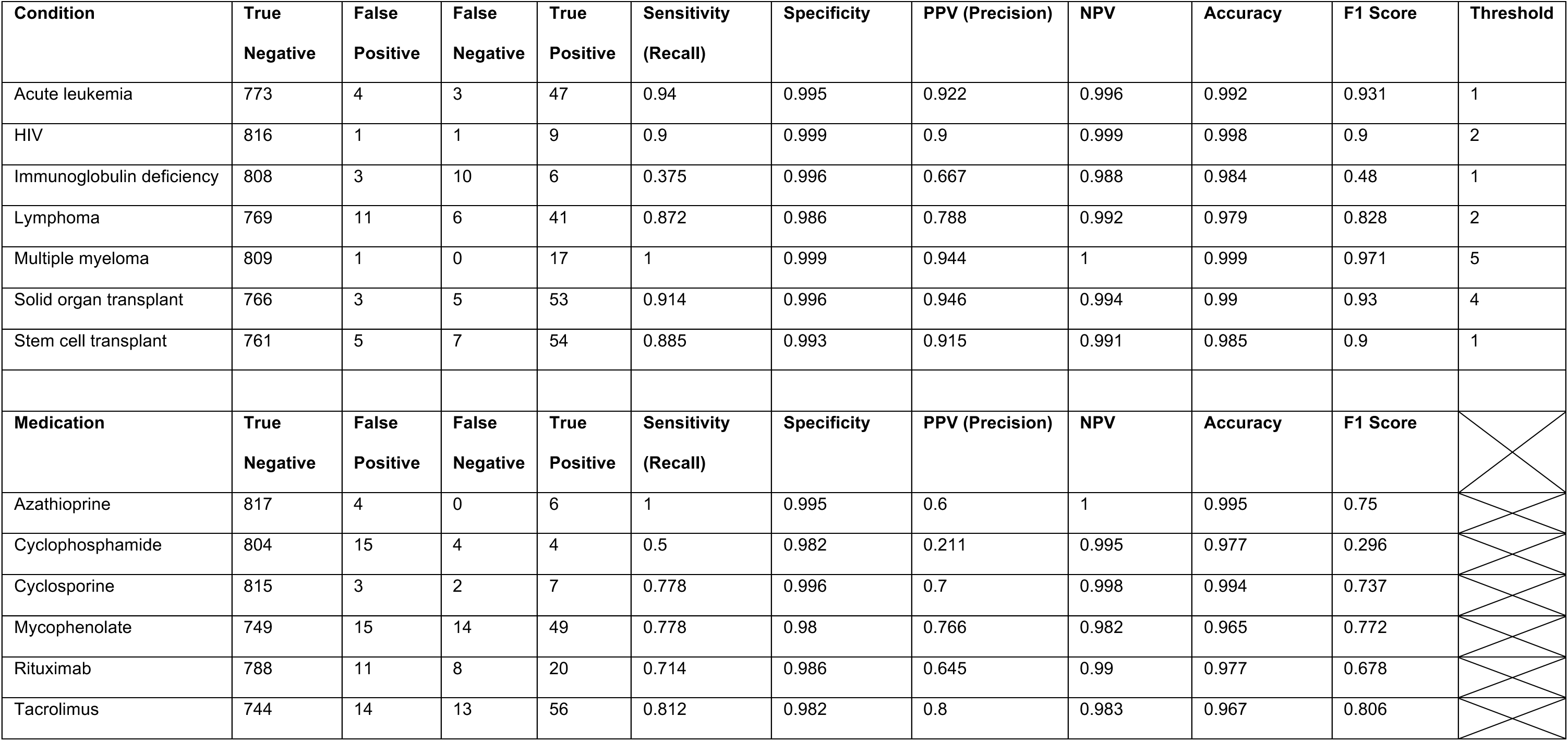
Performance of structured data in identifying immunosuppressive conditions and medications in the SCRIPT cohort. For conditions, metrics are shown at the code frequency threshold with the highest F1 score.

**Supplementary table 3.**
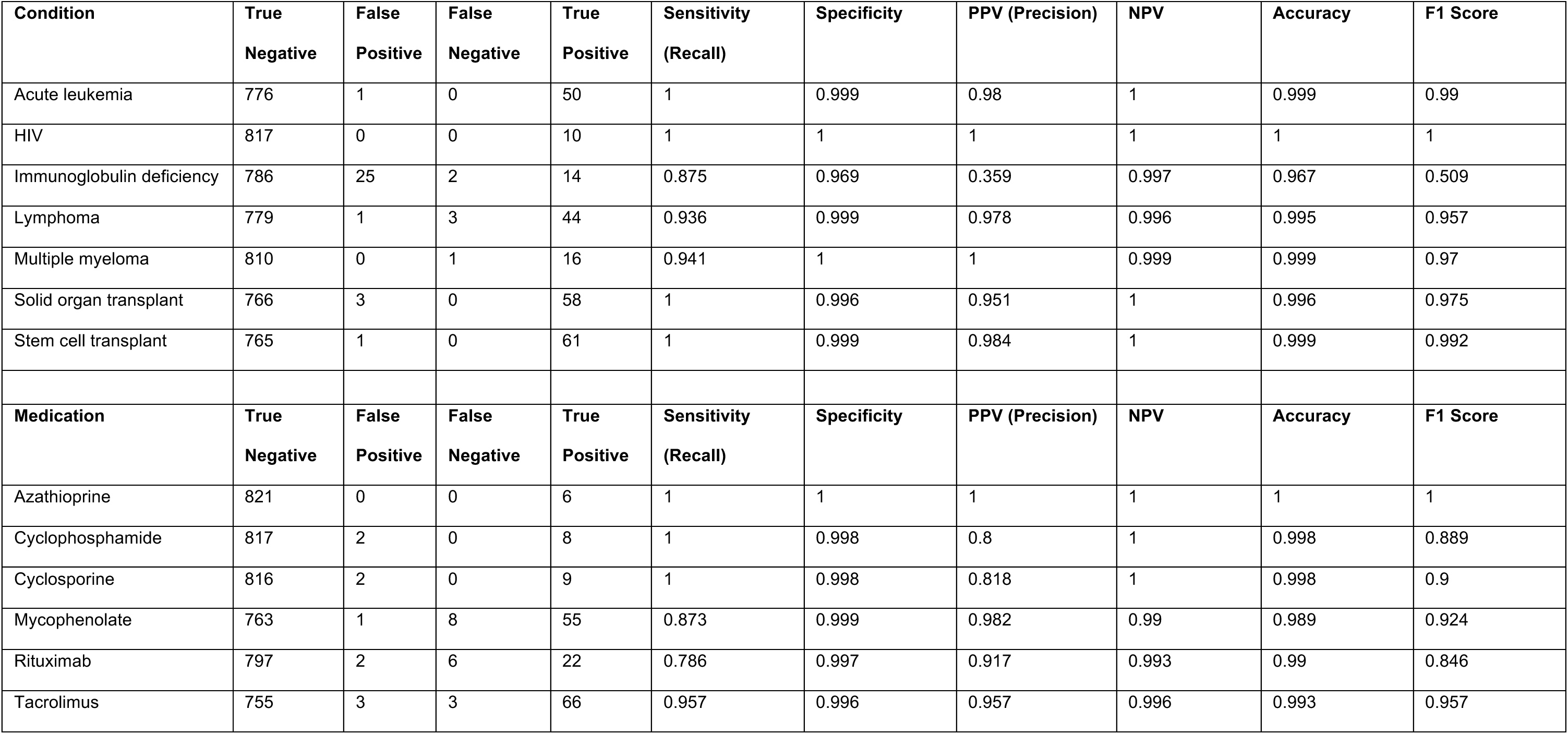
Performance of GPT-4o in identifying immunosuppressive conditions and medications in the SCRIPT cohort.

**Supplementary table 4.**
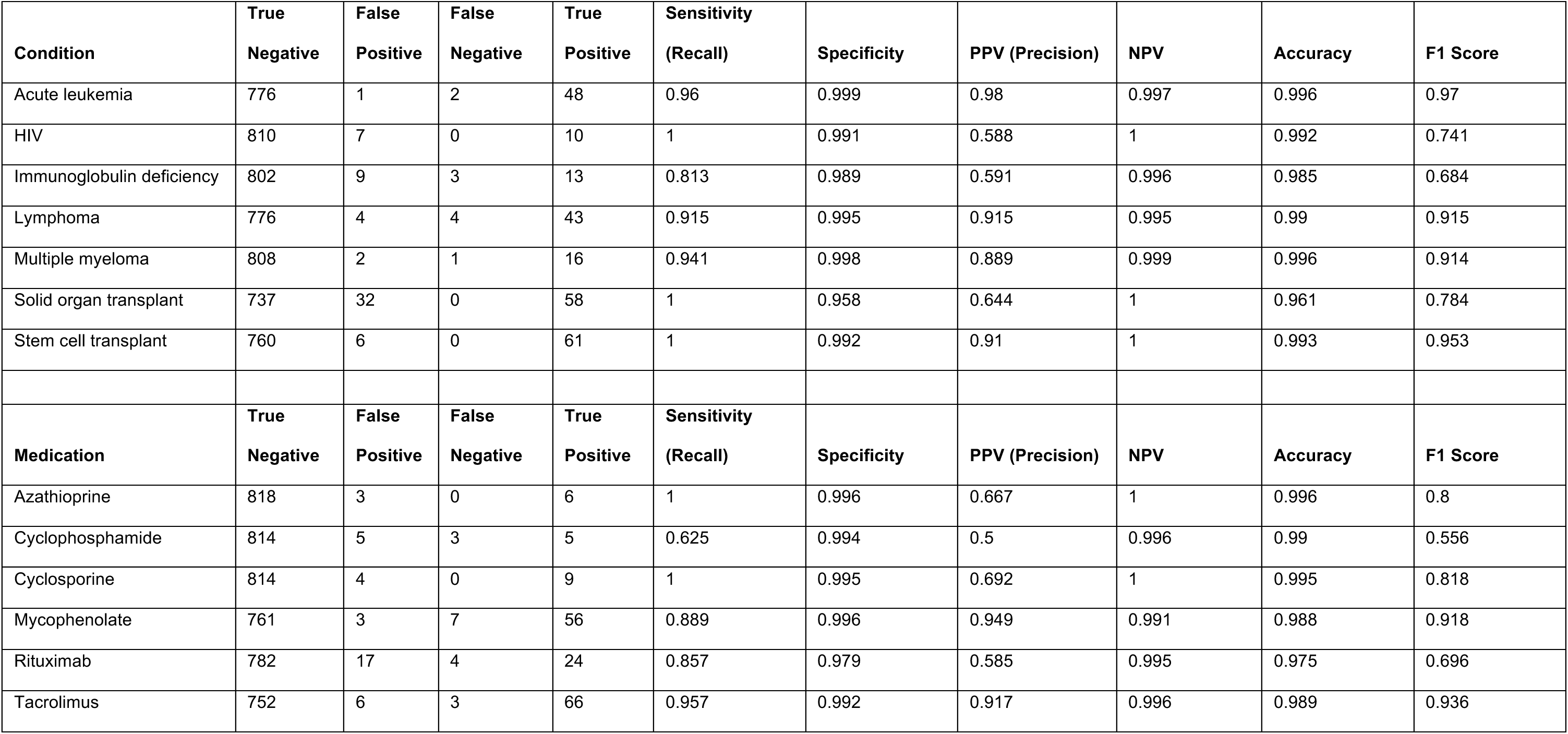
Performance of GPT-4o mini in identifying immunosuppressive conditions and medications in the SCRIPT cohort.

**Supplementary table 5.**
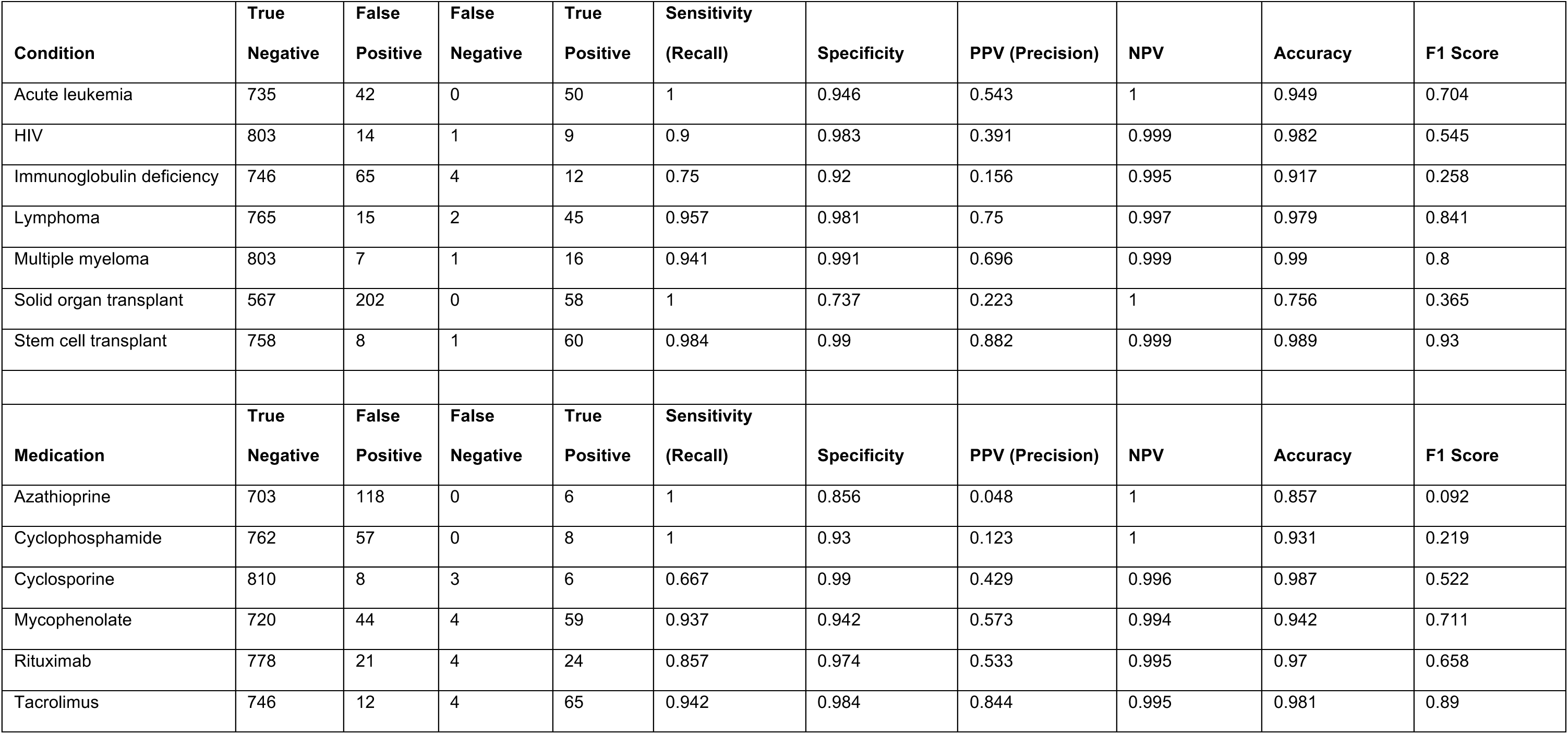
Performance of Llama 3.1 in identifying immunosuppressive conditions and medications in the SCRIPT cohort.

**Supplementary table 6.**
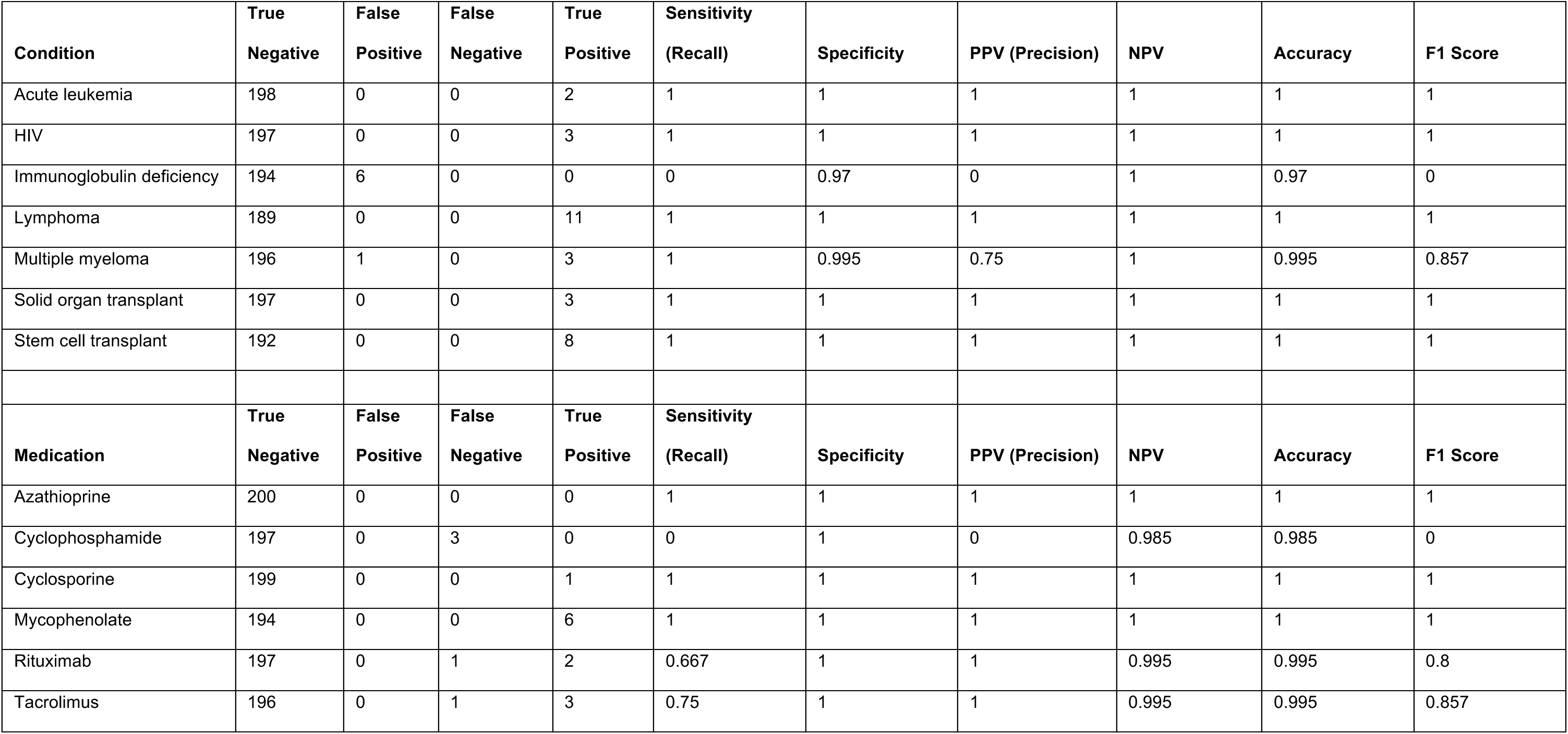
Performance of GPT-4o in identifying immunosuppressive conditions and medications in the MIMIC-III cohort.

**Supplementary table 7.**
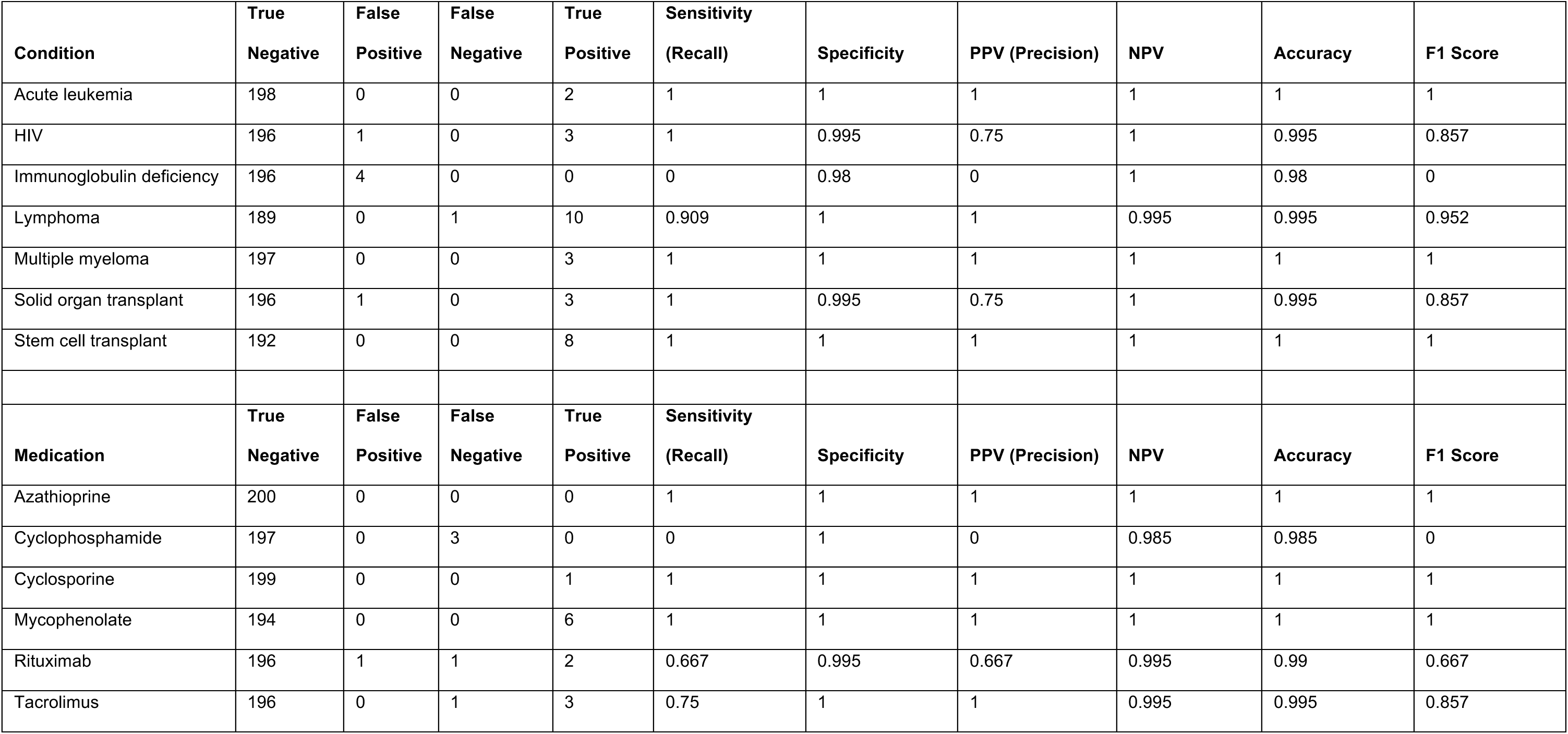
Performance of GPT-4o mini in identifying immunosuppressive conditions and medications in the MIMIC-III cohort.

**Supplementary table 8.**
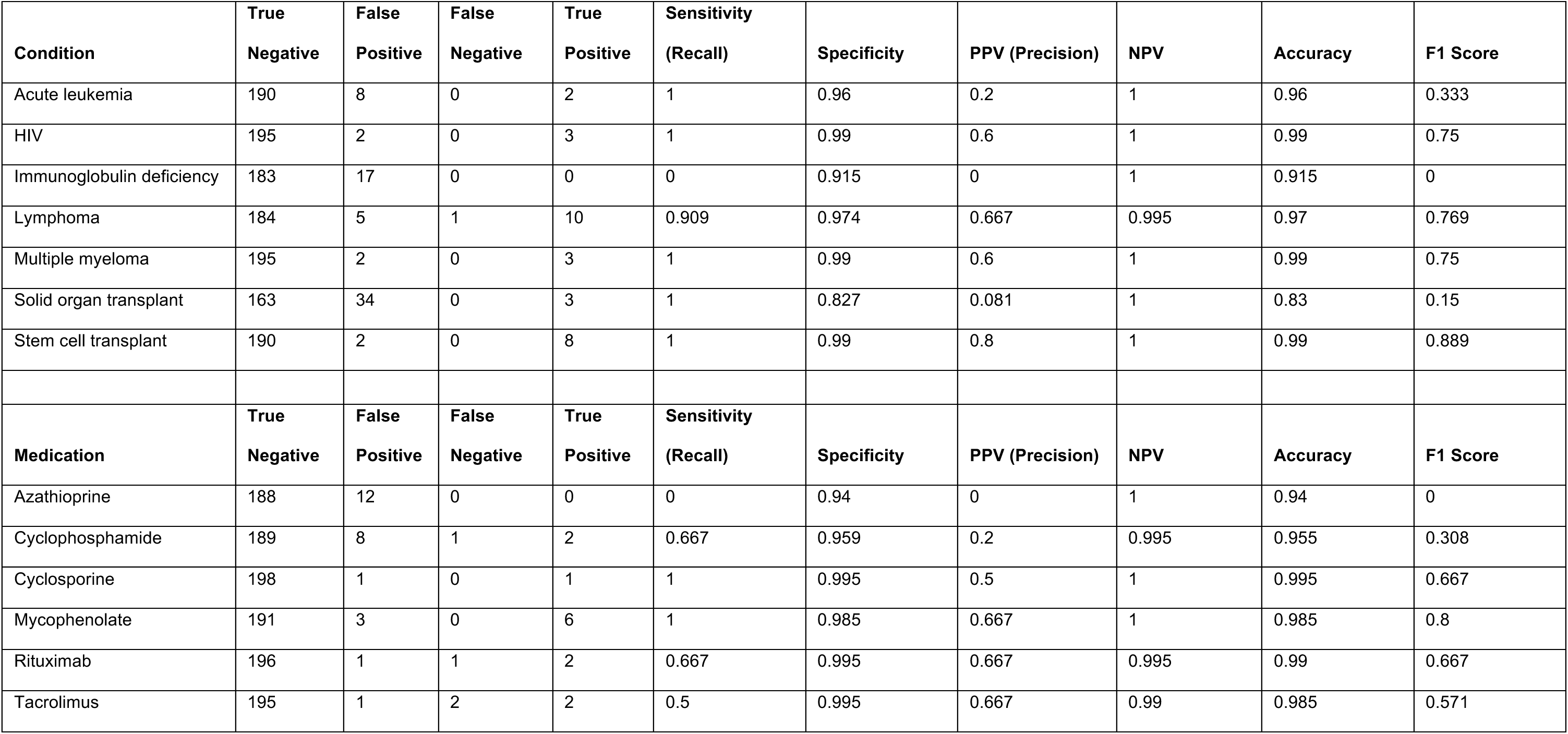
Performance of Llama 3.1 in identifying immunosuppressive conditions and medications in the MIMIC-III cohort.

**Supplementary table 9.**
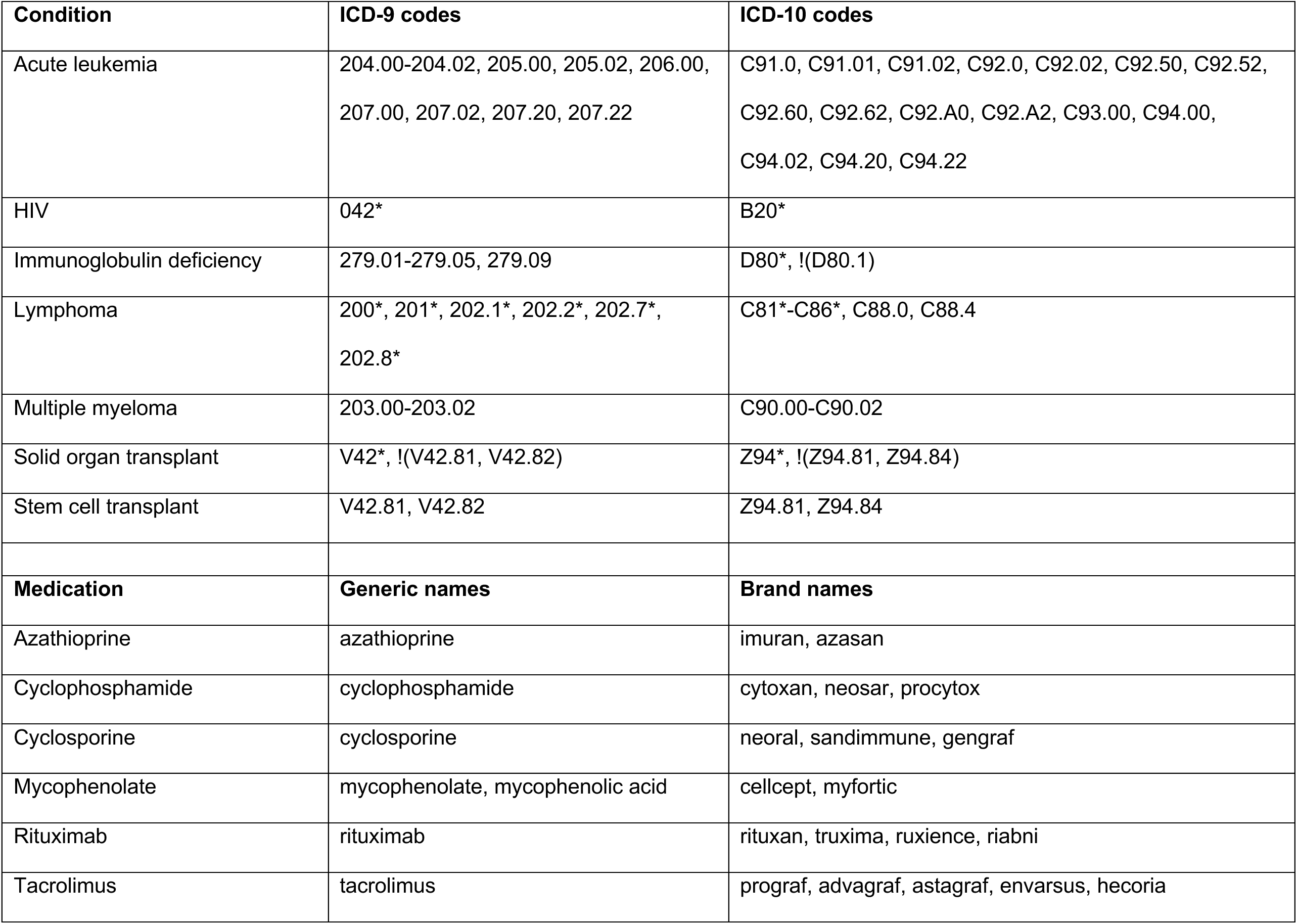
ICD-9/ICD-10 diagnosis codes and medication generic/brand names used for structured data predictions.

## Notes

### Author Declarations

IRB of Northwestern University gave ethical approval for this work (STU00204868).

