## Supplementary material for "Large language models outperform traditional structured data-based approaches in identifying immunosuppressed patients"

### Supplementary information

**Supplementary table 1.** Summary of F1 scores for all identification methods of all immunosuppressive conditions and medications in both the SCRIPT and MIMIC-III cohorts.

| **SCRIPT** | | | | |
|---|---|---|---|---|
| **Condition** | **Structured Data Peak F1 Score** | **GPT-4o F1 Score** | **GPT-4o mini F1 Score** | **Llama 3.1 F1 Score** |
| Acute leukemia | 0.931 | 0.99 | 0.97 | 0.704 |
| HIV | 0.9 | 1 | 0.741 | 0.545 |
| Immunoglobulin deficiency | 0.48 | 0.509 | 0.684 | 0.258 |
| Lymphoma | 0.828 | 0.957 | 0.915 | 0.841 |
| Multiple myeloma | 0.971 | 0.97 | 0.914 | 0.8 |
| Solid organ transplant | 0.93 | 0.975 | 0.784 | 0.365 |
| Stem cell transplant | 0.9 | 0.992 | 0.953 | 0.93 |
| **Medication** | **Structured Data Peak F1 Score** | **GPT-4o F1 Score** | **GPT-4o mini F1 Score** | **Llama 3.1 F1 Score** |
| Azathioprine | 0.75 | 1 | 0.8 | 0.092 |
| Cyclophosphamide | 0.296 | 0.889 | 0.556 | 0.219 |
| Cyclosporine | 0.737 | 0.9 | 0.818 | 0.522 |
| Mycophenolate | 0.772 | 0.924 | 0.918 | 0.711 |
| Rituximab | 0.678 | 0.846 | 0.696 | 0.658 |
| Tacrolimus | 0.806 | 0.957 | 0.936 | 0.89 |
| **MIMIC-III** | | | | |
| **Condition** | | **GPT-4o F1 Score** | **GPT-4o mini F1 Score** | **Llama 3.1 F1 Score** |
| Acute leukemia | | 1 | 1 | 0.333 |
| HIV | | 1 | 0.857 | 0.75 |
| Immunoglobulin deficiency | | 0 | 0 | 0 |

| | | | | |
|---|---|---|---|---|
| Lymphoma | | 1 | 0.952 | 0.769 |
| Multiple myeloma | | 0.857 | 1 | 0.75 |
| Solid organ transplant | | 1 | 0.857 | 0.15 |
| Stem cell transplant | | 1 | 1 | 0.889 |
| **Medication** | | **GPT-4o F1 Score** | **GPT-4o mini F1 Score** | **Llama 3.1 F1 Score** |
| Azathioprine | | 1 | 1 | 0 |
| Cyclophosphamide | | 0 | 0 | 0.308 |
| Cyclosporine | | 1 | 1 | 0.667 |
| Mycophenolate | | 1 | 1 | 0.8 |
| Rituximab | | 0.8 | 0.667 | 0.667 |
| Tacrolimus | | 0.857 | 0.857 | 0.571 |

**Supplementary table 2.** Performance of structured data in identifying immunosuppressive conditions and medications in the SCRIPT cohort. For conditions, metrics are shown at the code frequency threshold with the highest F1 score.

| Condition | True Negative | False Positive | False Negative | True Positive | Sensitivity (Recall) | Specificity | PPV (Precision) | NPV | Accuracy | F1 Score | Threshold |
|---|---|---|---|---|---|---|---|---|---|---|---|
| Acute leukemia | 773 | 4 | 3 | 47 | 0.94 | 0.995 | 0.922 | 0.996 | 0.992 | 0.931 | 1 |
| HIV | 816 | 1 | 1 | 9 | 0.9 | 0.999 | 0.9 | 0.999 | 0.998 | 0.9 | 2 |
| Immunoglobulin deficiency | 808 | 3 | 10 | 6 | 0.375 | 0.996 | 0.667 | 0.988 | 0.984 | 0.48 | 1 |
| Lymphoma | 769 | 11 | 6 | 41 | 0.872 | 0.986 | 0.788 | 0.992 | 0.979 | 0.828 | 2 |
| Multiple myeloma | 809 | 1 | 0 | 17 | 1 | 0.999 | 0.944 | 1 | 0.999 | 0.971 | 5 |
| Solid organ transplant | 766 | 3 | 5 | 53 | 0.914 | 0.996 | 0.946 | 0.994 | 0.99 | 0.93 | 4 |
| Stem cell transplant | 761 | 5 | 7 | 54 | 0.885 | 0.993 | 0.915 | 0.991 | 0.985 | 0.9 | 1 |
| **Medication** | **True Negative** | **False Positive** | **False Negative** | **True Positive** | **Sensitivity (Recall)** | **Specificity** | **PPV (Precision)** | **NPV** | **Accuracy** | **F1 Score** | |
| Azathioprine | 817 | 4 | 0 | 6 | 1 | 0.995 | 0.6 | 1 | 0.995 | 0.75 | |
| Cyclophosphamide | 804 | 15 | 4 | 4 | 0.5 | 0.982 | 0.211 | 0.995 | 0.977 | 0.296 | |
| Cyclosporine | 815 | 3 | 2 | 7 | 0.778 | 0.996 | 0.7 | 0.998 | 0.994 | 0.737 | |
| Mycophenolate | 749 | 15 | 14 | 49 | 0.778 | 0.98 | 0.766 | 0.982 | 0.965 | 0.772 | |
| Rituximab | 788 | 11 | 8 | 20 | 0.714 | 0.986 | 0.645 | 0.99 | 0.977 | 0.678 | |
| Tacrolimus | 744 | 14 | 13 | 56 | 0.812 | 0.982 | 0.8 | 0.983 | 0.967 | 0.806 | |

**Supplementary table 3.** Performance of GPT-4o in identifying immunosuppressive conditions and medications in the SCRIPT cohort.

| Condition | True Negative | False Positive | False Negative | True Positive | Sensitivity (Recall) | Specificity | PPV (Precision) | NPV | Accuracy | F1 Score |
|---|---|---|---|---|---|---|---|---|---|---|
| Acute leukemia | 776 | 1 | 0 | 50 | 1 | 0.999 | 0.98 | 1 | 0.999 | 0.99 |
| HIV | 817 | 0 | 0 | 10 | 1 | 1 | 1 | 1 | 1 | 1 |
| Immunoglobulin deficiency | 786 | 25 | 2 | 14 | 0.875 | 0.969 | 0.359 | 0.997 | 0.967 | 0.509 |
| Lymphoma | 779 | 1 | 3 | 44 | 0.936 | 0.999 | 0.978 | 0.996 | 0.995 | 0.957 |
| Multiple myeloma | 810 | 0 | 1 | 16 | 0.941 | 1 | 1 | 0.999 | 0.999 | 0.97 |
| Solid organ transplant | 766 | 3 | 0 | 58 | 1 | 0.996 | 0.951 | 1 | 0.996 | 0.975 |
| Stem cell transplant | 765 | 1 | 0 | 61 | 1 | 0.999 | 0.984 | 1 | 0.999 | 0.992 |
| **Medication** | **True Negative** | **False Positive** | **False Negative** | **True Positive** | **Sensitivity (Recall)** | **Specificity** | **PPV (Precision)** | **NPV** | **Accuracy** | **F1 Score** |
| Azathioprine | 821 | 0 | 0 | 6 | 1 | 1 | 1 | 1 | 1 | 1 |
| Cyclophosphamide | 817 | 2 | 0 | 8 | 1 | 0.998 | 0.8 | 1 | 0.998 | 0.889 |
| Cyclosporine | 816 | 2 | 0 | 9 | 1 | 0.998 | 0.818 | 1 | 0.998 | 0.9 |
| Mycophenolate | 763 | 1 | 8 | 55 | 0.873 | 0.999 | 0.982 | 0.99 | 0.989 | 0.924 |
| Rituximab | 797 | 2 | 6 | 22 | 0.786 | 0.997 | 0.917 | 0.993 | 0.99 | 0.846 |
| Tacrolimus | 755 | 3 | 3 | 66 | 0.957 | 0.996 | 0.957 | 0.996 | 0.993 | 0.957 |

**Supplementary table 4.** Performance of GPT-4o mini in identifying immunosuppressive conditions and medications in the SCRIPT cohort.

| Condition | True Negative | False Positive | False Negative | True Positive | Sensitivity (Recall) | Specificity | PPV (Precision) | NPV | Accuracy | F1 Score |
|---|---|---|---|---|---|---|---|---|---|---|
| Acute leukemia | 776 | 1 | 2 | 48 | 0.96 | 0.999 | 0.98 | 0.997 | 0.996 | 0.97 |
| HIV | 810 | 7 | 0 | 10 | 1 | 0.991 | 0.588 | 1 | 0.992 | 0.741 |
| Immunoglobulin deficiency | 802 | 9 | 3 | 13 | 0.813 | 0.989 | 0.591 | 0.996 | 0.985 | 0.684 |
| Lymphoma | 776 | 4 | 4 | 43 | 0.915 | 0.995 | 0.915 | 0.995 | 0.99 | 0.915 |
| Multiple myeloma | 808 | 2 | 1 | 16 | 0.941 | 0.998 | 0.889 | 0.999 | 0.996 | 0.914 |
| Solid organ transplant | 737 | 32 | 0 | 58 | 1 | 0.958 | 0.644 | 1 | 0.961 | 0.784 |
| Stem cell transplant | 760 | 6 | 0 | 61 | 1 | 0.992 | 0.91 | 1 | 0.993 | 0.953 |
| **Medication** | **True Negative** | **False Positive** | **False Negative** | **True Positive** | **Sensitivity (Recall)** | **Specificity** | **PPV (Precision)** | **NPV** | **Accuracy** | **F1 Score** |
| Azathioprine | 818 | 3 | 0 | 6 | 1 | 0.996 | 0.667 | 1 | 0.996 | 0.8 |
| Cyclophosphamide | 814 | 5 | 3 | 5 | 0.625 | 0.994 | 0.5 | 0.996 | 0.99 | 0.556 |
| Cyclosporine | 814 | 4 | 0 | 9 | 1 | 0.995 | 0.692 | 1 | 0.995 | 0.818 |
| Mycophenolate | 761 | 3 | 7 | 56 | 0.889 | 0.996 | 0.949 | 0.991 | 0.988 | 0.918 |
| Rituximab | 782 | 17 | 4 | 24 | 0.857 | 0.979 | 0.585 | 0.995 | 0.975 | 0.696 |
| Tacrolimus | 752 | 6 | 3 | 66 | 0.957 | 0.992 | 0.917 | 0.996 | 0.989 | 0.936 |

**Supplementary table 5.** Performance of Llama 3.1 in identifying immunosuppressive conditions and medications in the SCRIPT cohort.

| **Condition** | **True Negative** | **False Positive** | **False Negative** | **True Positive** | **Sensitivity (Recall)** | **Specificity** | **PPV (Precision)** | **NPV** | **Accuracy** | **F1 Score** |
|---|---|---|---|---|---|---|---|---|---|---|
| Acute leukemia | 735 | 42 | 0 | 50 | 1 | 0.946 | 0.543 | 1 | 0.949 | 0.704 |
| HIV | 803 | 14 | 1 | 9 | 0.9 | 0.983 | 0.391 | 0.999 | 0.982 | 0.545 |
| Immunoglobulin deficiency | 746 | 65 | 4 | 12 | 0.75 | 0.92 | 0.156 | 0.995 | 0.917 | 0.258 |
| Lymphoma | 765 | 15 | 2 | 45 | 0.957 | 0.981 | 0.75 | 0.997 | 0.979 | 0.841 |
| Multiple myeloma | 803 | 7 | 1 | 16 | 0.941 | 0.991 | 0.696 | 0.999 | 0.99 | 0.8 |
| Solid organ transplant | 567 | 202 | 0 | 58 | 1 | 0.737 | 0.223 | 1 | 0.756 | 0.365 |
| Stem cell transplant | 758 | 8 | 1 | 60 | 0.984 | 0.99 | 0.882 | 0.999 | 0.989 | 0.93 |
| **Medication** | **True Negative** | **False Positive** | **False Negative** | **True Positive** | **Sensitivity (Recall)** | **Specificity** | **PPV (Precision)** | **NPV** | **Accuracy** | **F1 Score** |
| Azathioprine | 703 | 118 | 0 | 6 | 1 | 0.856 | 0.048 | 1 | 0.857 | 0.092 |
| Cyclophosphamide | 762 | 57 | 0 | 8 | 1 | 0.93 | 0.123 | 1 | 0.931 | 0.219 |
| Cyclosporine | 810 | 8 | 3 | 6 | 0.667 | 0.99 | 0.429 | 0.996 | 0.987 | 0.522 |
| Mycophenolate | 720 | 44 | 4 | 59 | 0.937 | 0.942 | 0.573 | 0.994 | 0.942 | 0.711 |
| Rituximab | 778 | 21 | 4 | 24 | 0.857 | 0.974 | 0.533 | 0.995 | 0.97 | 0.658 |
| Tacrolimus | 746 | 12 | 4 | 65 | 0.942 | 0.984 | 0.844 | 0.995 | 0.981 | 0.89 |

**Supplementary table 6.** Performance of GPT-4o in identifying immunosuppressive conditions and medications in the MIMIC-III cohort.

| Condition | True Negative | False Positive | False Negative | True Positive | Sensitivity (Recall) | Specificity | PPV (Precision) | NPV | Accuracy | F1 Score |
|---|---|---|---|---|---|---|---|---|---|---|
| Acute leukemia | 198 | 0 | 0 | 2 | 1 | 1 | 1 | 1 | 1 | 1 |
| HIV | 197 | 0 | 0 | 3 | 1 | 1 | 1 | 1 | 1 | 1 |
| Immunoglobulin deficiency | 194 | 6 | 0 | 0 | 0 | 0.97 | 0 | 1 | 0.97 | 0 |
| Lymphoma | 189 | 0 | 0 | 11 | 1 | 1 | 1 | 1 | 1 | 1 |
| Multiple myeloma | 196 | 1 | 0 | 3 | 1 | 0.995 | 0.75 | 1 | 0.995 | 0.857 |
| Solid organ transplant | 197 | 0 | 0 | 3 | 1 | 1 | 1 | 1 | 1 | 1 |
| Stem cell transplant | 192 | 0 | 0 | 8 | 1 | 1 | 1 | 1 | 1 | 1 |
| **Medication** | **True Negative** | **False Positive** | **False Negative** | **True Positive** | **Sensitivity (Recall)** | **Specificity** | **PPV (Precision)** | **NPV** | **Accuracy** | **F1 Score** |
| Azathioprine | 200 | 0 | 0 | 0 | 1 | 1 | 1 | 1 | 1 | 1 |
| Cyclophosphamide | 197 | 0 | 3 | 0 | 0 | 1 | 0 | 0.985 | 0.985 | 0 |
| Cyclosporine | 199 | 0 | 0 | 1 | 1 | 1 | 1 | 1 | 1 | 1 |
| Mycophenolate | 194 | 0 | 0 | 6 | 1 | 1 | 1 | 1 | 1 | 1 |
| Rituximab | 197 | 0 | 1 | 2 | 0.667 | 1 | 1 | 0.995 | 0.995 | 0.8 |
| Tacrolimus | 196 | 0 | 1 | 3 | 0.75 | 1 | 1 | 0.995 | 0.995 | 0.857 |

**Supplementary table 7.** Performance of GPT-4o mini in identifying immunosuppressive conditions and medications in the MIMIC-III cohort.

| Condition | True Negative | False Positive | False Negative | True Positive | Sensitivity (Recall) | Specificity | PPV (Precision) | NPV | Accuracy | F1 Score |
|---|---|---|---|---|---|---|---|---|---|---|
| Acute leukemia | 198 | 0 | 0 | 2 | 1 | 1 | 1 | 1 | 1 | 1 |
| HIV | 196 | 1 | 0 | 3 | 1 | 0.995 | 0.75 | 1 | 0.995 | 0.857 |
| Immunoglobulin deficiency | 196 | 4 | 0 | 0 | 0 | 0.98 | 0 | 1 | 0.98 | 0 |
| Lymphoma | 189 | 0 | 1 | 10 | 0.909 | 1 | 1 | 0.995 | 0.995 | 0.952 |
| Multiple myeloma | 197 | 0 | 0 | 3 | 1 | 1 | 1 | 1 | 1 | 1 |
| Solid organ transplant | 196 | 1 | 0 | 3 | 1 | 0.995 | 0.75 | 1 | 0.995 | 0.857 |
| Stem cell transplant | 192 | 0 | 0 | 8 | 1 | 1 | 1 | 1 | 1 | 1 |
| **Medication** | **True Negative** | **False Positive** | **False Negative** | **True Positive** | **Sensitivity (Recall)** | **Specificity** | **PPV (Precision)** | **NPV** | **Accuracy** | **F1 Score** |
| Azathioprine | 200 | 0 | 0 | 0 | 1 | 1 | 1 | 1 | 1 | 1 |
| Cyclophosphamide | 197 | 0 | 3 | 0 | 0 | 1 | 0 | 0.985 | 0.985 | 0 |
| Cyclosporine | 199 | 0 | 0 | 1 | 1 | 1 | 1 | 1 | 1 | 1 |
| Mycophenolate | 194 | 0 | 0 | 6 | 1 | 1 | 1 | 1 | 1 | 1 |
| Rituximab | 196 | 1 | 1 | 2 | 0.667 | 0.995 | 0.667 | 0.995 | 0.99 | 0.667 |
| Tacrolimus | 196 | 0 | 1 | 3 | 0.75 | 1 | 1 | 0.995 | 0.995 | 0.857 |

**Supplementary table 8.** Performance of Llama 3.1 in identifying immunosuppressive conditions and medications in the MIMIC-III cohort.

| **Condition** | **True Negative** | **False Positive** | **False Negative** | **True Positive** | **Sensitivity (Recall)** | **Specificity** | **PPV (Precision)** | **NPV** | **Accuracy** | **F1 Score** |
|---|---|---|---|---|---|---|---|---|---|---|
| Acute leukemia | 190 | 8 | 0 | 2 | 1 | 0.96 | 0.2 | 1 | 0.96 | 0.333 |
| HIV | 195 | 2 | 0 | 3 | 1 | 0.99 | 0.6 | 1 | 0.99 | 0.75 |
| Immunoglobulin deficiency | 183 | 17 | 0 | 0 | 0 | 0.915 | 0 | 1 | 0.915 | 0 |
| Lymphoma | 184 | 5 | 1 | 10 | 0.909 | 0.974 | 0.667 | 0.995 | 0.97 | 0.769 |
| Multiple myeloma | 195 | 2 | 0 | 3 | 1 | 0.99 | 0.6 | 1 | 0.99 | 0.75 |
| Solid organ transplant | 163 | 34 | 0 | 3 | 1 | 0.827 | 0.081 | 1 | 0.83 | 0.15 |
| Stem cell transplant | 190 | 2 | 0 | 8 | 1 | 0.99 | 0.8 | 1 | 0.99 | 0.889 |
| **Medication** | **True Negative** | **False Positive** | **False Negative** | **True Positive** | **Sensitivity (Recall)** | **Specificity** | **PPV (Precision)** | **NPV** | **Accuracy** | **F1 Score** |
| Azathioprine | 188 | 12 | 0 | 0 | 0 | 0.94 | 0 | 1 | 0.94 | 0 |
| Cyclophosphamide | 189 | 8 | 1 | 2 | 0.667 | 0.959 | 0.2 | 0.995 | 0.955 | 0.308 |
| Cyclosporine | 198 | 1 | 0 | 1 | 1 | 0.995 | 0.5 | 1 | 0.995 | 0.667 |
| Mycophenolate | 191 | 3 | 0 | 6 | 1 | 0.985 | 0.667 | 1 | 0.985 | 0.8 |
| Rituximab | 196 | 1 | 1 | 2 | 0.667 | 0.995 | 0.667 | 0.995 | 0.99 | 0.667 |
| Tacrolimus | 195 | 1 | 2 | 2 | 0.5 | 0.995 | 0.667 | 0.99 | 0.985 | 0.571 |

**Supplementary table 9.** ICD-9/ICD-10 diagnosis codes and medication generic/brand names used for structured data predictions.

| **Condition** | **ICD-9 codes** | **ICD-10 codes** |
|---|---|---|
| Acute leukemia | 204.00-204.02, 205.00, 205.02, 206.00, 207.00, 207.02, 207.20, 207.22 | C91.0, C91.01, C91.02, C92.0, C92.02, C92.50, C92.52, C92.60, C92.62, C92.A0, C92.A2, C93.00, C94.00, C94.02, C94.20, C94.22 |
| HIV | 042* | B20* |
| Immunoglobulin deficiency | 279.01-279.05, 279.09 | D80*, !(D80.1) |
| Lymphoma | 200*, 201*, 202.1*, 202.2*, 202.7*, 202.8* | C81*-C86*, C88.0, C88.4 |
| Multiple myeloma | 203.00-203.02 | C90.00-C90.02 |
| Solid organ transplant | V42*, !(V42.81, V42.82) | Z94*, !(Z94.81, Z94.84) |
| Stem cell transplant | V42.81, V42.82 | Z94.81, Z94.84 |
| **Medication** | **Generic names** | **Brand names** |
| Azathioprine | azathioprine | imuran, azasan |
| Cyclophosphamide | cyclophosphamide | cytoxan, neosar, procytox |
| Cyclosporine | cyclosporine | neoral, sandimmune, gengraf |
| Mycophenolate | mycophenolate, mycophenolic acid | cellcept, myfortic |
| Rituximab | rituximab | rituxan, truxima, ruxience, riabni |
| Tacrolimus | tacrolimus | prograf, advagraf, astagraf, envarsus, hecoria |
